# Government Responses Matter: Predicting Covid-19 cases in US using an empirical Bayesian time series framework

**DOI:** 10.1101/2020.03.28.20044578

**Authors:** Ziyue Liu, Wensheng Guo

## Abstract

Since the Covid-19 outbreak, researchers have been predicting how the epidemic will evolve, especially the number in each country, through using parametric extrapolations based on the history. In reality, the epidemic progressing in a particular country depends largely on its policy responses and interventions. Since the outbreaks in some countries are earlier than United States, the prediction of US cases can benefit from incorporating the similarity in their trajectories. We propose an empirical Bayesian time series framework to predict US cases using different countries as prior reference. The resultant forecast is based on observed US data and prior information from the reference country while accounting for different population sizes. When Italy is used as prior in the prediction, which the US data resemble the most, the cases in the US will exceed 300,000 by the beginning of April unless strong measures are adopted.

---

When facing an epidemic, people and government of a country may underestimate its seriousness in the beginning but will eventually step up their responses. Hence the case numbers tend to increase exponentially in the early stage, while the trends will gradually bend and plateau. Therefore, similarities in the case number trajectories can be observed in different countries, though the timing and severity can differ substantially due to different responses. Figure 1 displays the trajectories of total Covid-19 case numbers for China, S. Korea, Italy, France, Iran, Germany, Spain and USA using Johns Hopkins data. These countries have more days from time zero than US, where time zero is defined as first day with 100 or more (100+) cases as a heuristic but widely used choice^1^. The curve of South Korea increased rapidly early on but quickly bended and plateaued, for which S. Korea’s swift and deterministic policy responses are credited^2^. China exhibits similar but later flattening pattern, which agrees with its missing early intervention window, but later extreme lockdown policy implementation^3^. On the other hand, the cases in Italy and France have grown exponentially until recent days, which have partially been attributed to their late and weak policy responses^4^. The US trajectory is almost linear on the logarithm scale. While the US government is catching up with policies such as work/study from home, social distancing and self-quarantine, the effect has not seen in the trajectory.

**Figure 1.**
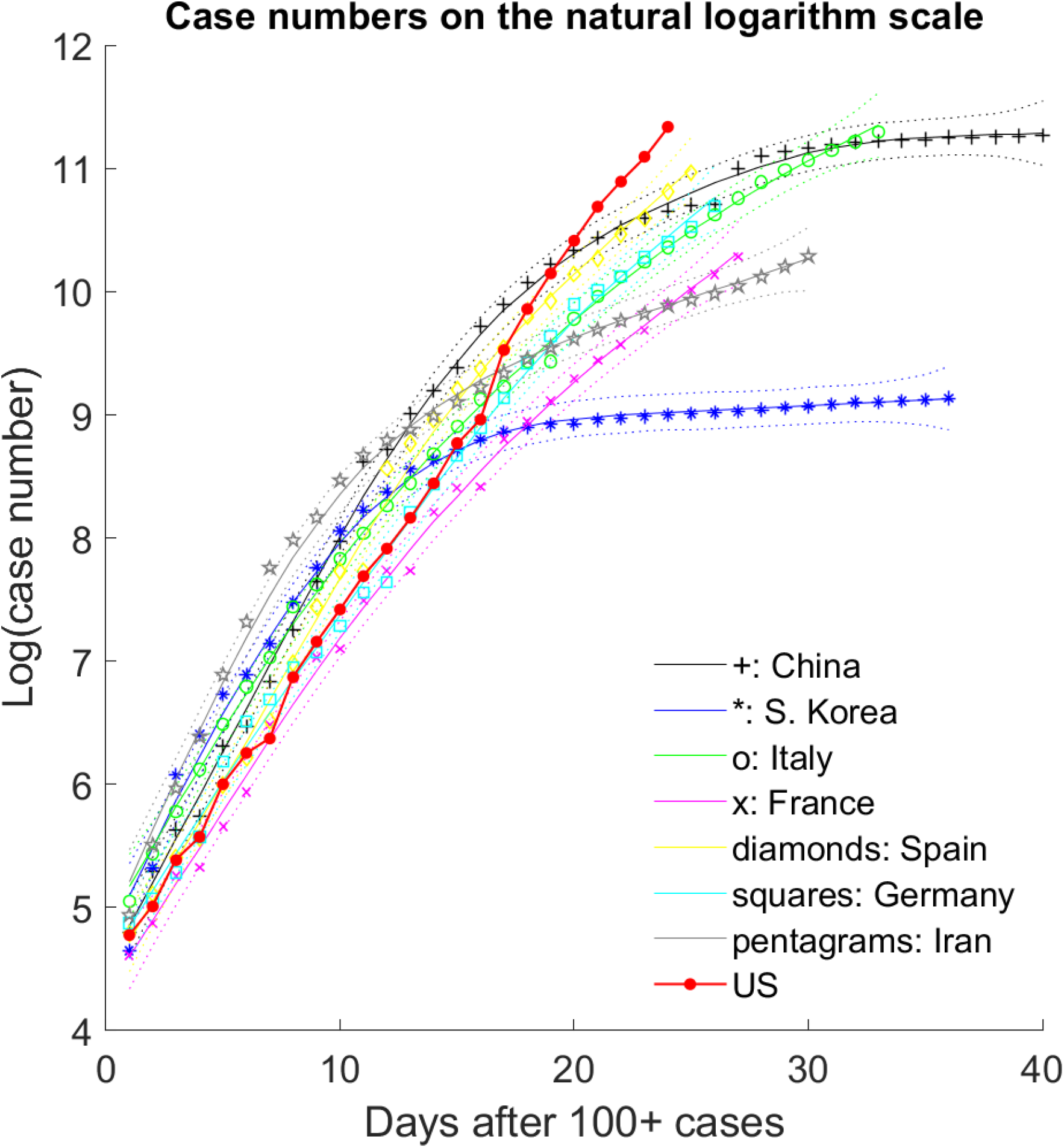
Cases numbers for China, S. Korea, Italy, France, Spain, Germany, Iran, and US on the natural logarithm scale. For the first seven countries, the raw data are shown as symbols, the smoothed trends as solid lines, and the 95% confidence intervals in dotted lines. For US, only the raw data are displayed.

Existing Covid-19 forecasting are extrapolations into the future time^5-11^. Their validity relies on the crucial but unrealistic assumption that the future trajectories are completely determined by the history. This by design cannot incorporate government responses yet to come. Not surprisingly, these predictions can be off the target. For example, Fanelli and Piazza^7^ predicted a maximum number of cases in Italy to be 15,000, where the real cases have already multipled. Batista^8^ predicted the pandemic should peak around Feb 9^th^, 2020, but it shows no sign of slowing down into late-March, 2020. Zheng et al^9^ predicted about 20,000 cases in South Korea, which is unlikely to happen given its current flat trend around 9,000. Models used in these forecasting are mainly the susceptible-infected-removed (SIR) models and its variants^5-8^. Others include state transition model^9^, parametric growth curve models such as logistic curves^10^, and auto regressive integrated moving average (ARIMA) models^11^.

## Proposed Methods

We propose an empirical Bayesian time series framework to forecast the US trajectory by utilizing the idea of internal time. Since the virus spread to different countries at different time, their trajectories are different in calendar time but comparable in internal time. We define time zero as the first day with 100 or more cases in a given country. We first model the trajectories of the eight countries by a functional mixed effects model^12^, where different countries shared a similar mean trajectory over time, and each country has its own random deviation curve. An additional scalar fixed effect parameter is incorporated to account for different population sizes on the natural logarithm scale. The estimated coefficient is 0.34, suggesting while the population size has some effects on the cases numbers, it is not fully proportion to the population. Both the population-average curve and random deviation curves are modeled by cubic splines. The model is then casted into state space model for computational efficiency and forecasting. The smoothing parameters and the variances are estimated through maximum likelihood.

Based on the estimated parameters using the eight countries, our next task is forecast the US cases while incorporate one of the countries as the prior information. This is done through constructing conditional state space model from the functional mixed effects model conditional on the observed data of the specified country^13^. By running the Kalman filter forward on the conditional state space model with the US time series data and into the future, the results are the posterior prediction incorporating both the prior information from the specific country and the observed US data. As the reference country is only specified as the prior, the posterior can be substantially different from the prior, suggesting strong deviation from the reference country.

In addition, the observed US data can be substantially different from posterior prediction, indicating that the US case are following a different trajectory because of different policy responses. More technical details are given in the Supplement.

### Data Analysis

The Johns Hopkins University CSSE data were downloaded from its GitHub repository (https://github.com/CSSEGISandData/COVID-19). We modeled the natural logarithms of the case numbers as the outcome. The data were then used for prediction using the proposed method. After the posterior means and variances were calculated and the 95% prediction intervals were constructed, they were taken exponential to transform back to the original scale. The whole data analysis from reading in the data to plotting the results took less than 10 seconds on personal computer with Intel® Core™i76600U CPU @ 2.60GHz, 2801Mhz, 2 Cores, 4 Logical Processors.

## Results

Results based on US data up to March 26^th^, 2020 are shown in Figure 2∼4. Two important observations can be made from these figures. There is no apparent slowing down yet for US trajectory based on either the observed trend or predicted trend. This indicates that US is still in its exponentially increasing phase in the near future.

**Figure 2.**
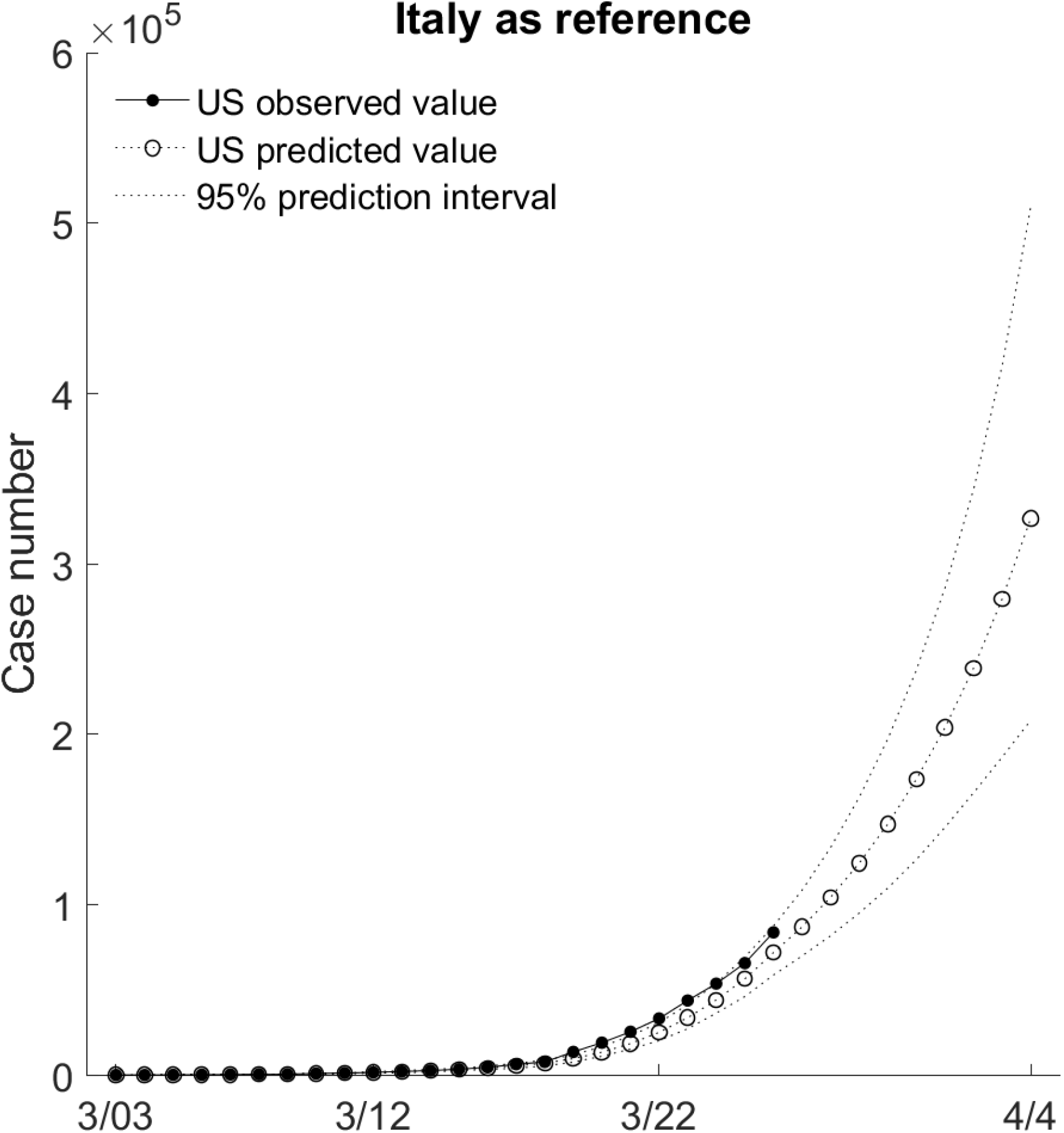
Prediction of the US trajectory using Italy as prior. The horizontal axis is based on US calendar time. On April 4^th^, 2020, the mean prediction value exceeds 300,000.

Figure 2 displays the results using Italy as prior. It shows that US and Italy have similar patterns and majority of the observed US data are in the 95% prediction intervals. This suggests that the trajectory in Italy serves as a good prior for the US prediction. Based on this prediction, on the next day as March 27^th^, 2020, US may have as many as 108,595 cases. In about 10 days, the US case number will exceed 300,000 around April 4^th^, 2020 shall the US policy responses have similarly effects as Italy.

Figure 3 displays the results using China as prior. It shows that the observed US case numbers are already higher than the predicted values. Even if the US policy responses have similar effect as China, US case numbers will exceed 150,000 around April 11^th^, 2020. The results using South Korea as prior are displayed in Figure 4. US case numbers are predicted to exceed 200,000 around April 6^th^, 2020. Since the observed US data are already well above the upper bound of the 95% prediction intervals, the data from China and South Korea are not good priors for the US prediction, suggesting that the situation in the US will be much worse than those in China and South Korea.

**Figure 3.**
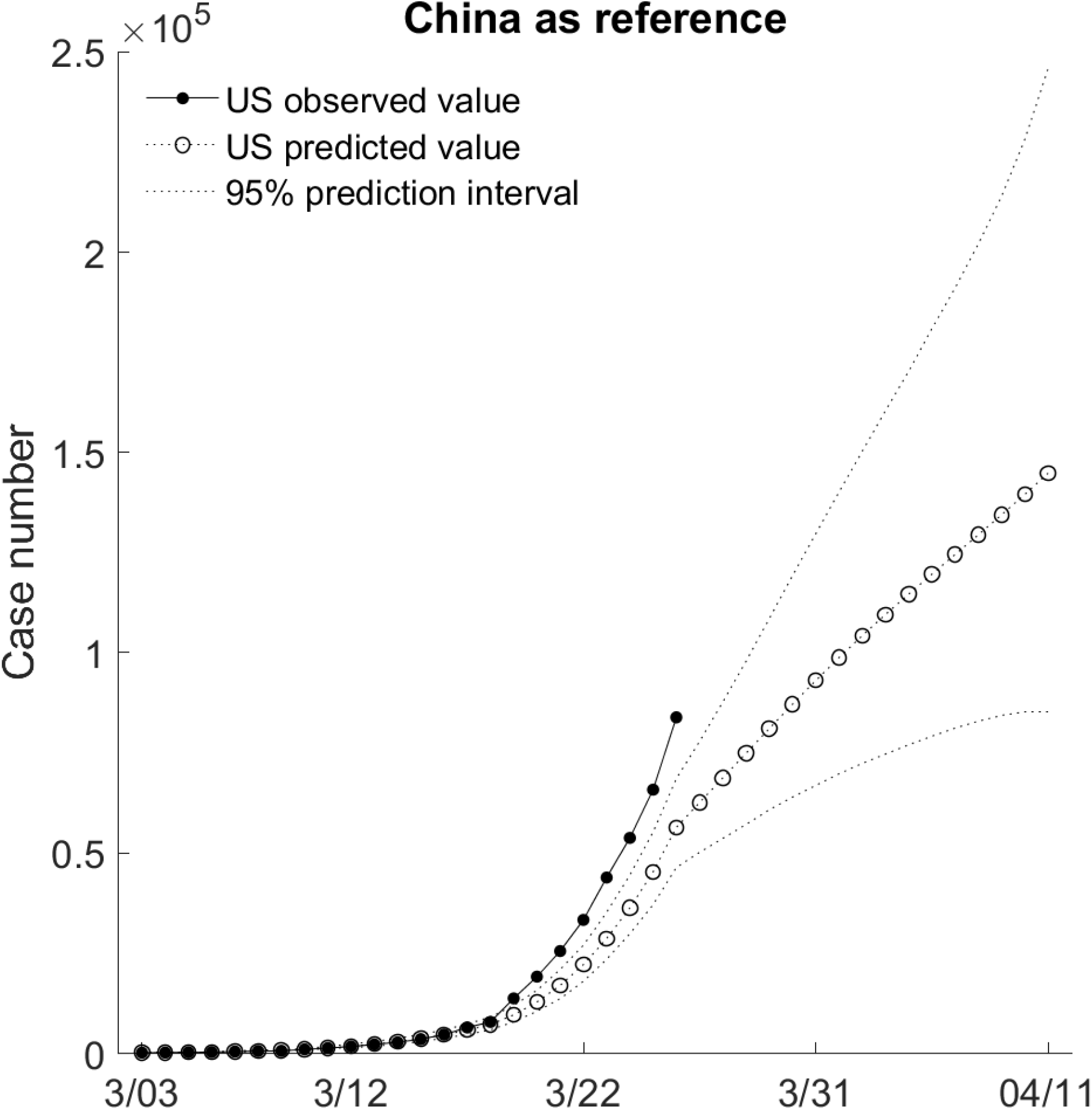
Prediction of the US trajectory using China as prior. The horizontal axis is based on US calendar time. On April 11^th^, 2020, the mean prediction value exceeds 150,000.

**Figure 4.**
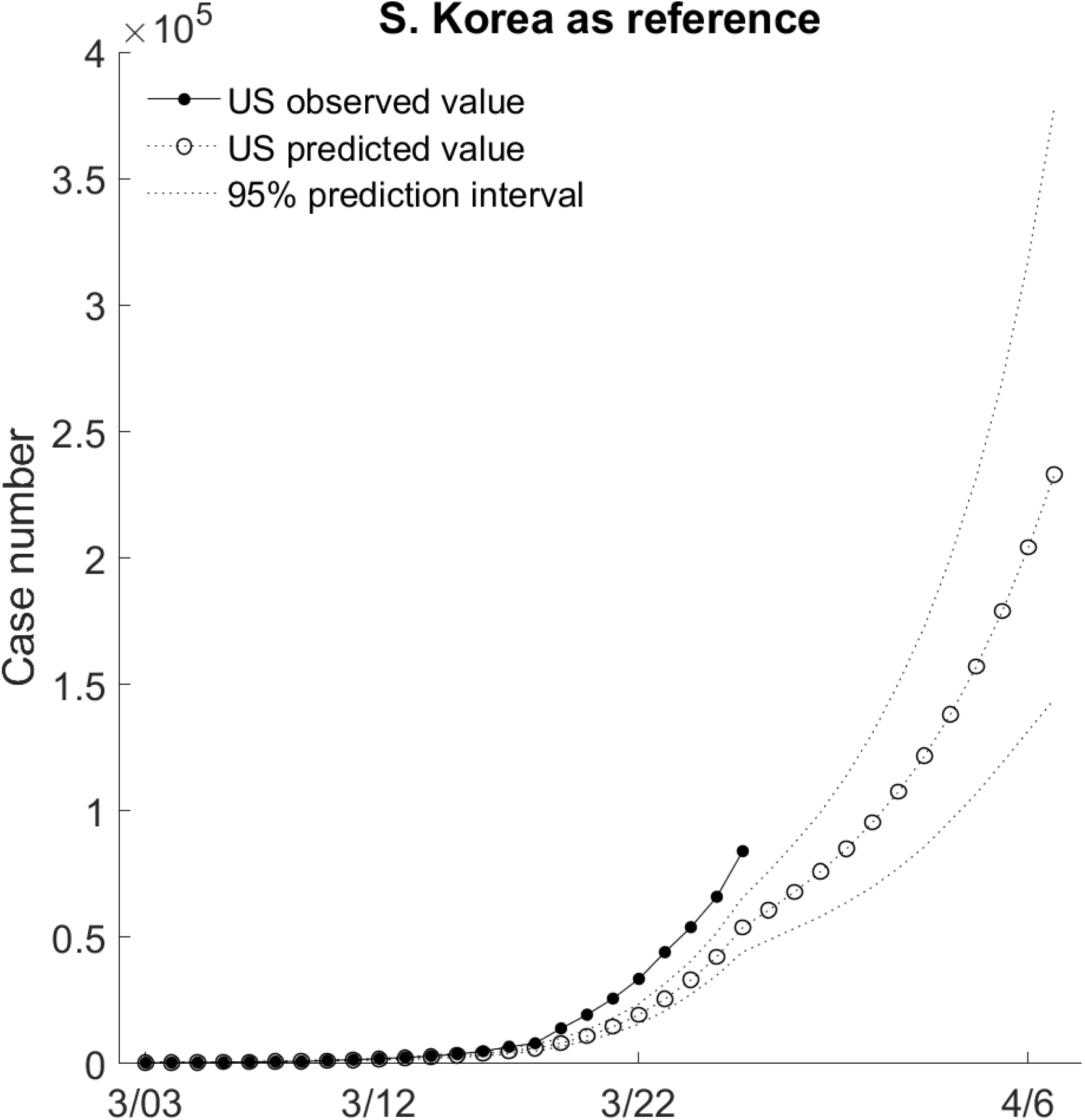
Prediction of the US trajectory using S. Korea as prior. The horizontal axis is based on US calendar time. On April 6^th^, the mean prediction value exceeds 200,000.

## Conclusion

We have proposed a new prediction method for predicting total COVID-19 cases of US by incorporating the information from other countries. While we demonstrated our method in predicting US cases, our method can be used for predicting state-by-state data as well as hospital-by-hospital data. Our prediction intervals are much smaller than most exiting methods due to the additional information from the reference country. We show that the current trajectory in US is most similar to that in Italy. The stronger response from Italy has led to slowing down of the spread in the last few days, while the effect of social distancing in the US has not shown in the observed data.

It is well-known that there are serious under-reporting or under-detection of cases in various countries and under-reporting rates may be very different across counties. This can contribute to substantial differences in the trajectories. With the advance of testing techniques, more and more people are tested in the US. This may also explain why the reported cases in the US are substantially higher than other countries in the same stages.

## Data Availability

Data and Matlab code for this manuscript are included as a supplement.

## Supplement

### Functional mixed effects model

Let *N*_*ij*_ be the number of total COVID-19 cases for the *i*^*th*^ country on the *j*^*th*^ day, where day 1 is defined as the first day with 100 or more cases. We model the natural logarithm of *N*_*ij*_, *y*_*ij*_ = log(*N*_*ij*_), by a functional mixed effects model^1^ as

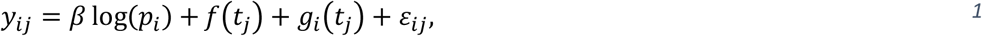

Where *p*_*i*_ is the population size in the unit of millions, *β* is the fixed effect slope for log(*p*_*i*_), *f*(*t*_*j*_) is the functional fixed effects, *g*_*i*_(*t*_*j*_) is the functional random effects, and 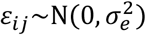 is the error term. We model *f*(*t*_*j*_) by a cubic smoothing spline with the state space representation as

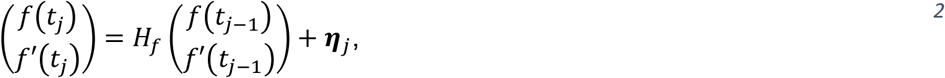

where *f*′(*t*_*j*_) is the first derivative with respect to time. The state transition matrix 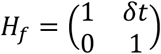 with *δt* is the time interval between two points with the overall time range scaled to [0,1]. The state innovation vector ***η***_*j*_∼N(**0**, Σ_*f*_) with 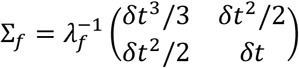 and *λ* is the smoothing parameter. The state vector 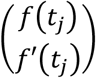 is initialized at time zero as 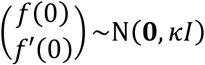 with *κ* → ∞ and *I* is the identity matrix. We model *g*_*i*_(*t*_*j*_) similarly but using a sine function in the *W*_0_ space with the state space representation^2^ as

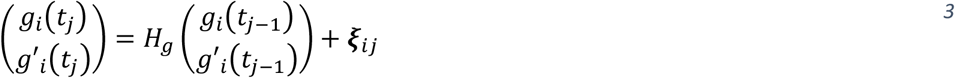

with time rescaled to [0, 0.5]. The state transition matrix 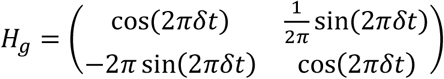.

The state innovation vector ***ξ***_*ij*_∼***N***(**0**, Σ_*g*_), with

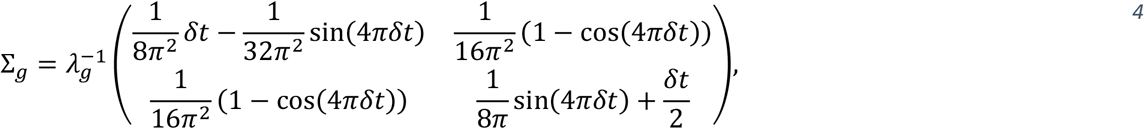

and *λ*_*g*_ is the smoothing parameter. The state vector 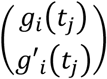 is initialized at time zero as 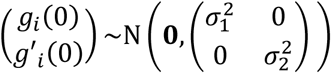.

### Parameter estimation

Data from eight countries (China, S. Korea, Italy, France, Iran, Germany, Spain and USA) were used. Let *n*_*i*_ denote the number of observations for subject *i* = 1, …, *k*, ***y***_*i*_ the corresponding observed data vector, ***t***_*i*_ the time vector, ***f***(***t***_*i*_) the vector of fixed functional effects evaluated at ***t***_*i*_, ***g***_*i*_(***t***_*i*_) the functional random effect, *f*”(*t*) and *g*”_*i*_(*t*) the second derivatives with respect to time, and ***y*** the overall observed data vector. The following penalized log-likelihood^3^ was maximized to estimate the parameter vector 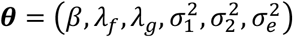

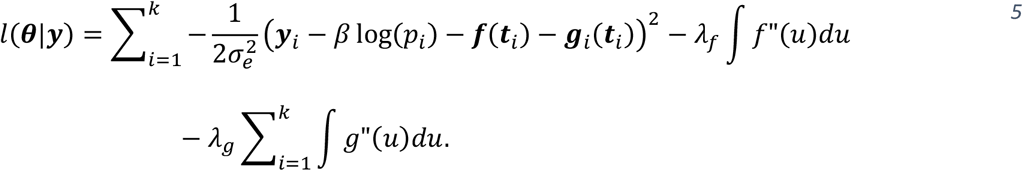

The maximum likelihood parameter estimates were 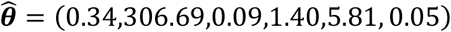. We adopt an empirical Bayes approach such that these parameters are treated as known in the following steps.

### Construction of the conditional SSM

For the *i*^*th*^ reference country, the conditional SSM was constructed on the state vectors dimension of six as 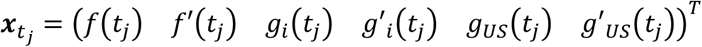, where subscript ‘US’ denote US-specific component. The working data are 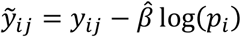. The observation matrix is *F* = (1 0 1 0 0 0), the state transition matrix is *H* = block diagonal (*H*_*f*_, *H*_*g*_, *H*_*g*_), the state innovation vector 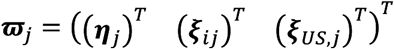 distributed as ***ϖ*** ∼N(**0**, Σ_*j*_) where Σ_*j*_ = block diagonal (Σ_*f*_, Σ_*g*_, Σ_*g*_). The working SSM is

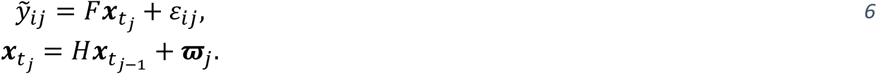

Let ***m***(**·**) denote the mean and *W*(**·**) the variance of 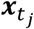. The conditional SSM was constructed in three steps using the dynamic state space models^4^.

Step 1. The forward filtering: for *j* = 1, …, *n*_*i*_

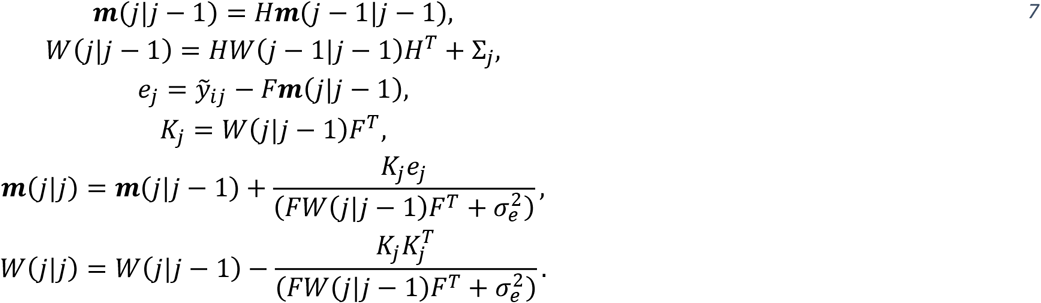

Step 2. The backward smoothing: at *j* = *n*_*i*_ let ***m***(*j*|*n*_*i*_) = ***m***(*n*_*i*_|*n*_*i*_) and *W*(*j*|*n*_*i*_) = *W*(*n*_*i*_|*n*_*i*_), for *j* = *n*_*i*_ − 1, …, 0

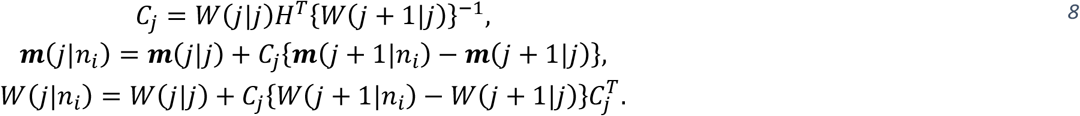

Step 3. Construction step: 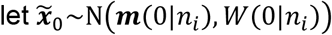, for *j* = 1, …, *n*_*i*_

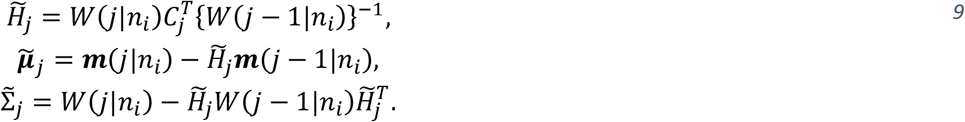

where 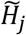 is the state transition matrix, 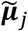 is the mean vector and 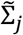 is the variance matrix of the state innovations.

### Predict US trajectory

The US trajectory was predicted by running the *n*_*US*_ observations 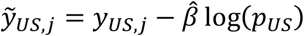 through the conditional SSM constructed above with an observation matrix *F* = (1 0 0 0 1 0). For *j* = 1, …, *n*_*US*_, the first two equations in (7) generated the one-step ahead prediction, while the last two equations generate the filtered values. Further running the model for *j* = *n*_*US*_ + 1, …, *n*_*i*_ generated predictions into the future time for US. 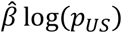 was then added back to recover the effects of population size.

